# Abnormally high exertional breathlessness predicts mortality in people referred for incremental cycle exercise testing

**DOI:** 10.1101/2024.03.31.24305143

**Authors:** Viktor Elmberg, Xingwu Zhou, Thomas Lindow, Kristofer Hedman, Andrei Malinovschi, Hayley Lewthwaite, Dennis Jensen, Lars Brudin, Magnus Ekström

**Affiliations:** Respiratory Medicine, Allergology and Palliative Medicine, Department of Clinical Sciences in Lund, Lund University, Lund, Sweden; Department of Clinical Physiology, Blekinge Hospital, Karlskrona, Sweden; Department of Medial Sciences, Clinical Physiology, Uppsala University, Uppsala, Sweden; Department of Clinical Physiology, Research and Development, Växjö Central Hospital, Region Kronoberg, and Clinical Sciences, Lund University, Sweden; Faculty of Medicine and Health, University of Sydney, Sydney, Australia; Department of Clinical Physiology in Linköping, and Department of Health, Medicine and Caring Sciences, Linköping University, Linköping, Sweden; Centre of Research Excellence Treatable Traits, College of Health, Medicine and Wellbeing, University of Newcastle, Newcastle, NSW, Australia; Asthma and Breathing Research Program, Hunter Medical Research Institute, Newcastle, NSW, Australia; Clinical Exercise and Respiratory Physiology Laboratory, Department of Kinesiology and Physical Education, McGill University, Montreal, Quebec, Canada; Research Institute of the McGill University Health Centre, Translational Research in Respiratory Diseases Program, Montreal, Quebec, Canada; Department of Clinical Physiology, Kalmar County Hospital, Kalmar,Sweden; Department of Medial Sciences, Respiratory-, Allergy- and Sleep Research, Uppsala University, Uppsala, Sweden

## Abstract

**Background:** Exertional breathlessness is a key symptom in cardiorespiratory disease and can be quantified using incremental exercise testing (IET), but its prognostic significance is unknown.

**Research question:** We evaluated the ability of abnormally high breathlessness intensity during IET to predict all-cause, respiratory, and cardiac mortality.

**Study Design and Methods:** Longitudinal cohort study of adults referred for cycle IET followed prospectively for mortality assessed using the Swedish National Causes of Death Registry. Abnormally high exertional breathlessness was defined as a breathlessness intensity response (Borg 0-10 scale) > the upper limit of normal (ULN) using published reference equations. Mortality was analyzed using multivariable Cox regression, unadjusted and adjusted for age, sex, and body mass index.

**Results:** Of the 13,506 people included (46% female, age 59±15 years), 2,867 (21%) had abnormally high breathlessness during IET. Over a median follow up of 8.0 years, 1,687 (12%) people died. No participant was lost to follow-up. Compared to those within normal predicted ranges, people with abnormally high exertional breathlessness had higher mortality from all causes (adjusted hazard ratio [aHR] 2.3, [95% confidence interval] 2.1-2.6), respiratory causes (aHR 5.2 [3.4-8.0]) and cardiac causes (aHR 3.0 [2.5-3.6]). Even among people with normal exercise capacity (defined as peak Watt ≥75% of predicted exercise capacity, n=10,284) those with abnormally high exertional breathlessness were at greater risk of all-cause mortality than people with exertional breathlessness within the normal predicted range (aHR 1.5 [1.2-1.8]).

**Interpretation:** Among people referred for cycle IET, abnormally high exertional breathlessness, quantified using healthy reference values, independently predicted all-cause, respiratory and cardiac mortality.

## INTRODUCTION

Exertional breathlessness is a cardinal symptom of cardiac and respiratory disease that often leads to a vicious cycle of reduced physical activity, with attendant further worsening of breathlessness, functional capacity, and quality of life (1, 2, 3, 4). Worse self-reported exertional breathlessness on daily life questionnaires has also been associated with higher risk of premature death (5, 6).

Breathlessness can be assessed in different ways, including questionnaires such as the modified Medical Research Council (mMRC) scale, which measures the self-reported limitation breathlessness imposes on daily life activities (7, 8). More standardized tests like incremental exercise testing (IET), with or without measurements of gas-exchange and ventilation (9, 10), are also commonly used and more optimal for measuring breathlessness as the symptom can be related to a power output (11). Reference equations derived from a Swedish cohort to predict the normal breathlessness response during cycle IET using the Borg Category-Ratio scale (Borg CR10) were recently published (12). Using these, the presence of abnormally high exertional breathlessness can be defined as a Borg CR10 intensity rating above the predicted upper limit of normal (ULN) at any given power output (W), expressed as a percentage of their predicted normal peak power output (%predWmax).

People referred for cardiac stress testing (IET, stress echocardiography and single-photon emission computerized tomography [SPECT]) because of breathlessness have, in a meta-analysis, been shown to have worse prognosis compared to those who were referred because of chest pain (13). Breathlessness as reason for termination of exercise during IET has also been shown to be a negative prognostic marker. In an earlier study, compared with people who stopped exercise due to “exhaustion” or “leg fatigue”, those who stopped due to breathlessness had worse survival (14). Self-reported breathlessness during stress testing has also been shown to be independently associated with both abnormal and high risk cardiac SPECT scans, both carrying an adverse prognosis (15). However, the prognostic implication of having an abnormally high breathlessness response to IET is unknown.

The primary aim of this study was to evaluate, in a longitudinal cohort of people referred for IET, whether abnormally high breathlessness intensity in relation to workload quantified using published reference equations (12), predicts all-cause, respiratory and/or cardiac mortality. Secondary aims were to (i) evaluate which %predW_max_ value during the IET best predicts mortality, and (ii) assess the prognostic significance of abnormally high exertional breathlessness intensity among people with normal and abnormally low peak exercise capacity (W_peak_).

## STUDY DESIGN AND METHODS

### Study design and participants

This was a longitudinal cohort study of adults aged 18 years or older who performed a standardized IET, according to the most used Swedish manual for exercise tests (16), at the Department of Clinical Physiology, Kalmar County Hospital, Sweden between 31^st^ May 2005 and 31^st^ October 2016 (Figure 1). The information contained in this database are such that it could potentially identify individual participants. The IET’s were performed for clinical reasons with the indication in most cases being suspect stable coronary syndrome (n=10,306), occupational risk evaluation (n=439), suspect arrhythmia (n=659) or determination of exercise capacity (n=562). This cohort has earlier been used as the basis for reference values for predicted normal peak power output (Wmax) (17) and exercise systolic blood pressure (18, 19), and breathlessness intensity response during IET (12).

**Figure 1.**
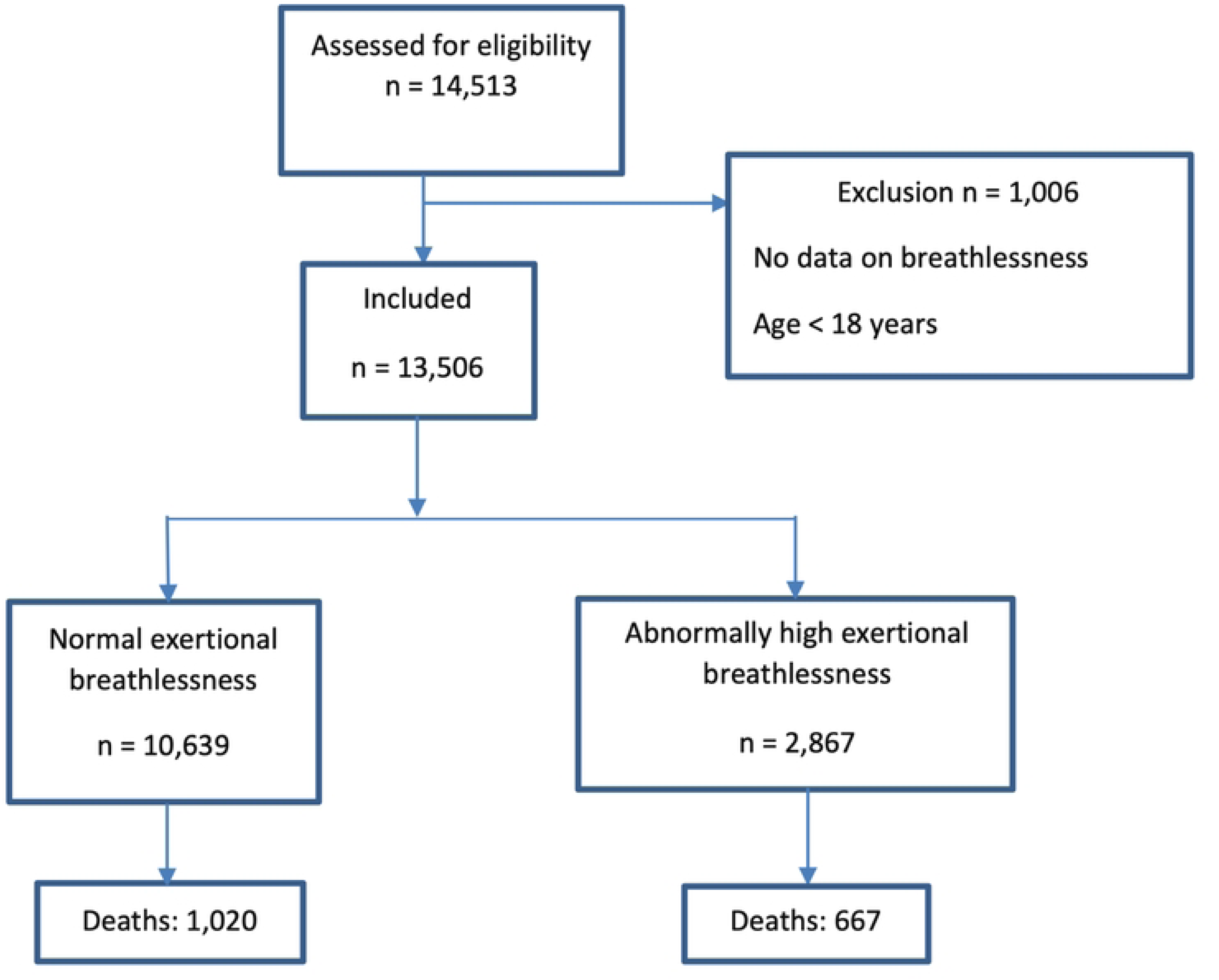
Flowchart illustrating the study design. Each individual was categorized as having normal or abnormally increased breathlessness during IET. The number of deaths in each group is shown.

Participants that did not rate their breathlessness at any time point during IET were excluded. The primary outcome mortality (all-cause, cardiac, and respiratory) was followed longitudinally using the mandatory Swedish Causes of Death Register (up until 30 April 2019), allowing for complete coverage. This registry contains the cause of mortality as listed on the medical death certificate recorded by the patient’s physician. The completeness and reliability of this registry is well established. (20). Comorbidity and hospital admissions data up until 30 December 2017 were available from the National Patient Register (21). All diagnoses from within 5 years preceding the IET were obtained and defined according to the International Classification of Diseases version 10 (ICD-10). These included for example chronic obstructive pulmonary disease (COPD), hypertension (HT), ischemic heart disease (IHD), heart failure (HF) and diabetes (DM). Data on mortality and hospitalization was accessed on the 15^th^ of May 2019.

### Ethical considerations

The study was conducted in accordance with the amended Declaration of Helsinki and was approved by the Regional Ethical Review Board in Linköping (DNr: 2018/141-31). As this was an observational study using data collected from IETs performed in clinical practice, individual participant consent was waived. The study is reported in accordance with the STrengthening the Reporting of OBservational studies in Epidemiology (STROBE) statement (22) and the TRIPOD statement (23).

### Exercise test protocol

The protocol for the standardized IET has been described elsewhere (17, 18, 24). In brief, all tests were performed using an electrically braked cycle ergometer (Rodby Inc, Karlskoga, Sweden). The initial power output (W) and ramp increment of 10, 15 or 20 W/min was selected depending on the participant’s predicted W_max_ (17), aiming at an exercise duration of 8-12 minutes (17, 18, 24). The effect of different power output increments on the achieved W_max_ is accounted for by the Swedish reference values for W_max_ (17, 25).

### Assessments

Before IET, 12-lead electrocardiogram (ECG), body mass and height were recorded. The Borg Rating of Perceived Exertion 6-20 scale (Borg RPE) for assessment of perceived exertion and Borg CR10 scale for assessment of chest pain and breathlessness (26) were explained before commencing the IET, including the scales anchor points. When assessing breathlessness, participants were invited to rate their intensity of perceived breathlessness on the Borg CR10 scale from 0, representing ‘no breathlessness’, to 10, representing ‘the most intense breathlessness that you’ve experienced or could imagine experiencing’ (26). During the IET, ECG was recorded continuously, whereas systolic blood pressure, RPE, breathlessness intensity and chest pain were measured every 2 minutes.

### Group allocation and breathlessness measurement

Abnormality of exertional breathlessness intensity was assessed at several %predW_max_ stages, specifically 25%, 50%, 75% and 100%. The stages were defined by intervals as a breathlessness rating might not be available at exactly 25% predW_max_. The corresponding intervals for the %predW_max_ stages were: 25% (15-35%), 50% (40-60%), 75% (65-85%) and 100% (90-110%). This method of allocation meant that the participants available for analysis decreased as the %predW_max_ stage increased. Each participant was also added to a fifth group (last measured) representing the exact %predW_max_ where the last breathlessness intensity rating was obtained close to or at W_peak_.

People were further divided into 4 groups according to exercise tolerance (%predW_max_) and exertional breathlessness response. This allowed for assessment of the impact of an abnormally high exertional breathlessness intensity response among people with normal or abnormally low exercise capacity on risk of death, as well as the interaction between abnormally low exercise capacity and abnormally high exertional breathlessness. Abnormally low exercise capacity was defined as having a W_peak_ <75% of predicted W_max_. The 4 groups were: Group 1 - normal exercise capacity + normal breathlessness intensity; Group 2 - normal exercise capacity + abnormally high breathlessness intensity; Group 3 - abnormally low exercise capacity + normal breathlessness intensity; and Group 4 - abnormally low exercise capacity + abnormally high breathlessness intensity.

### Statistical analyses

All analysis were conducted by R version 4.2.3 (R Core Team, 2023) (27). Characteristics were tabulated and compared using descriptive statistics. The normality or abnormality of exertional breathlessness intensity was determined using recently published Swedish reference equations (12). The equations predict the probability of rating each score on a Borg CR10 scale among healthy people based on sex, age, height (men) and %predW_max._ A p-value of < 0.05 is then used to define the ULN. Abnormally high exertional breathlessness was defined as having a breathlessness intensity rating > ULN. If a breathlessness intensity rating at any one or combination of %predW_max_ stages was abnormally high (defined as “all together”), the participant was identified as having abnormally high exertional breathlessness in the main analysis. The predictive value of every %predW_max_ stage was also analyzed independently.

Associations between the presence of abnormally high exertional breathlessness intensity and all-cause, cardiac, and respiratory mortality were analyzed using Cox proportional hazard regression expressed as hazard ratios (HRs) with 95% confidence intervals (CIs). Analyses were performed unadjusted and adjusted for age, sex, and body mass index (BMI). Kaplan-Meier plots were used to compare survival between people with and without abnormally high exertional breathlessness intensity. Association between abnormally high exertional breathlessness intensity and all-cause mortality were also analyzed separately corrected for %predW_max_ in addition to age, sex and BMI. Associations between abnormally high exertional breathlessness intensity and all-cause mortality was also analyzed for Groups 1, 2, 3 and 4, to account for the potentially confounded effect of exercise capacity on the association between abnormally high exertional breathlessness intensity and all-cause mortality.

Prognostic discriminative ability of all the models was analyzed using C-statistics, which is equal to the area under the curve (AUC) in receiver operator curve analysis.

## RESULTS

### Participants and breathlessness responses

A total of 13,506 participants were included (Figure 1). Their mean ± SD age was 59 ± 15 years (range 18–94) and 46% were female (Table 1). The most common reasons for cessation of exercise were leg fatigue (35%), general exertion (31%) and breathlessness (20%). Abnormally high exertional breathlessness intensity was present at any one or more exercise stages (all together) in 2,867 (21%) of the participants. The number of participants with normal, abnormally high, or missing breathlessness intensity ratings at each %predW_max_ stage during the IET is shown in supplemental Figure S1.

**Table 1.** Baseline participant characteristics stratified by the presence of a normal or abnormally high breathlessness intensity response during incremental cycle exercise testing.

| **Characteristic** | **Abnormally high exertional breathlessness intensity (Borg CR10 rating > ULN)**<br>**N = 2,867 (21%)** | **Normal exertional breathlessness intensity (Borg CR10 rating ≤ ULN)**<br>**N = 10,639 (79%)** |
| --- | --- | --- |
| Age | 61 ± 15 | 59 ± 15 |
| Male | 1,637 (57%) | 5,692 (54%) |
| Body height, cm | 172 ± 9 | 172 ± 10 |
| Body mass, kg | 81 ± 18 | 79 ± 15 |
| Body mass index, kg/m<sup>2</sup> | 27 ± 5 | 27 ± 4 |
| Systolic blood pressure supine, mmHg | 139 ± 26 | 138 ± 24 |
| Diastolic blood pressure supine, mmHg | 79 ± 11 | 79 ± 10 |
| Heart rate supine, beats/min | 76 ± 14 | 74 ± 13 |
| Comorbidities diagnosed before exercise test, n (%) |  |  |
| Chronic obstructive pulmonary disease | 193 (7%) | 129 (1%) |
| Heart failure | 157 (5%) | 115 (1%) |
| Diabetes mellitus | 328 (11%) | 519 (5%) |
| Ischemic heart disease | 459 (16%) | 944 (9%) |
| Hypertension | 591 (21%) | 1,262 (12%) |
| Malignancy | 340 (12%) | 952 (9%) |
| Valvular disease | 90 (3%) | 135 (1%) |
| Rheumatic disease | 107 (4%) | 252 (2%) |
| Renal failure | 96 (3%) | 99 (1%) |
| Self-reported use of medication, n (%) |  |  |
| Beta-blocker | 997 (35%) | 2,584 (24%) |
| Thrombocyte inhibitor | 823 (29%) | 2,062 (19%) |
| Statins | 741 (26%) | 1,949 (19%) |
| Warfarin/NOAC | 209 (7%) | 363 (34%) |
| ACEI/ARB | 1,589 (55%) | 2,640 (25%) |
| Calcium antagonist | 424 (15%) | 1,065 (10%) |
| Loop or Thiazide diuretics | 520 (18%) | 945 (9%) |
| Insulin | 208 (7%) | 343 (3%) |
| Other diabetic medication | 214 (8%) | 457 (4%) |
| Antidepressant | 366 (13%) | 886 (8%) |
| **Measures at peak exercise** |  |  |
| Power output, Watt | 126 ± 50 | 166 ± 60 |
| Power output, %predWmax | 71 ± 16 | 93 ± 17 |
| Breathlessness intensity, Borg CR10 scale units | 7.1± 1.7 | 5.9± 1.6 |
| Exertion, Borg RPE 6-20 scale units | 17.4 ± 1.2 | 17.1 ± 1.1 |
| Chest pain, Borg CR10 scale units | 0.8 ± 1.7 | 0.5 ± 1.3 |
| Heart rate, beats/min | 139 ± 27 | 152 ± 24 |
| Heart rate, % of predicted* | 88 ± 14 | 94 ± 12 |

Data presented as means ± standard deviation unless indicated otherwise. \*Predicted peak heart rate was calculated as 220-age (years). *Abbreviations:* Borg CR10 = Borg Category-ratio scale (0-10); %predWmax = W % of predicted Wmax; Borg RPE = Borg rating of perceived exertion scale (6-20); W = Watt; NOAC = nonwarfarin oral anticoagulant; ACEI = angiotensin-converting enzyme inhibitor; ARB = angiotensin receptor blocker.

Sex distribution and anthropometric measurements were similar between people with normal and abnormally high exertional breathlessness intensity (Table 1). People with abnormally high exertional breathlessness had a higher prevalence of medical conditions, including COPD, HF, DM, and HT (Table 1) as well as a lower W_peak_ (126 ± 50 W *vs.* 166 ± 60 W, p ≤ 0.001) and %predW_max_ (71 ± 16 % *vs.* 93 ± 17 %, p ≤ 0.001).

Mean peak breathlessness intensity (7.1 ± 1.7 vs. 5.9 ± 1.6 Borg CR10 scale units, p ≤ 0.001) and perceived exertion (17.4 ± 1.2 vs 17.1 ± 1.1 Borg RPE units, p ≤ 0.001) were higher in the group of participants with compared to without abnormally high exertional breathlessness intensity. Peak chest pain was also slightly higher in the group with abnormally high exertional breathlessness: 0.8 ± 1.7 vs 0.5 ± 1.3 Borg CR10 scale units, p ≤ 0.001.

### Mortality

No person was lost to follow-up. During a median follow-up of 8.0 years (Interquartile Range [IQR] 5.3-10.1; range 1.3-13.8), 1,687 (12%) participants died (Figure 2 a). Compared to those with normal exertional breathlessness intensity, participants with abnormally high breathlessness intensity had higher all-cause mortality: HR = 2.3 (2.1-2.6) adjusted for age, sex, and BMI (Table 2). When %predW_max_ was added to the adjusted analysis, the association between abnormally high exertional breathlessness and all-cause mortality was HR = 1.2 (1.0-1.3). The association between abnormally high exertional breathlessness and respiratory mortality was HR = 5.2 (3.4-8.0), whereas the association between abnormally high exertional breathlessness and cardiac mortality was HR = 3.0 (2.5-3.6) (Table 2, Figure 2 b/c). As illustrated in Figure 3, abnormally high exertional breathlessness was associated with a greater risk of all-cause mortality among people with both normal and abnormally low exercise capacity.

**Figure 2.**
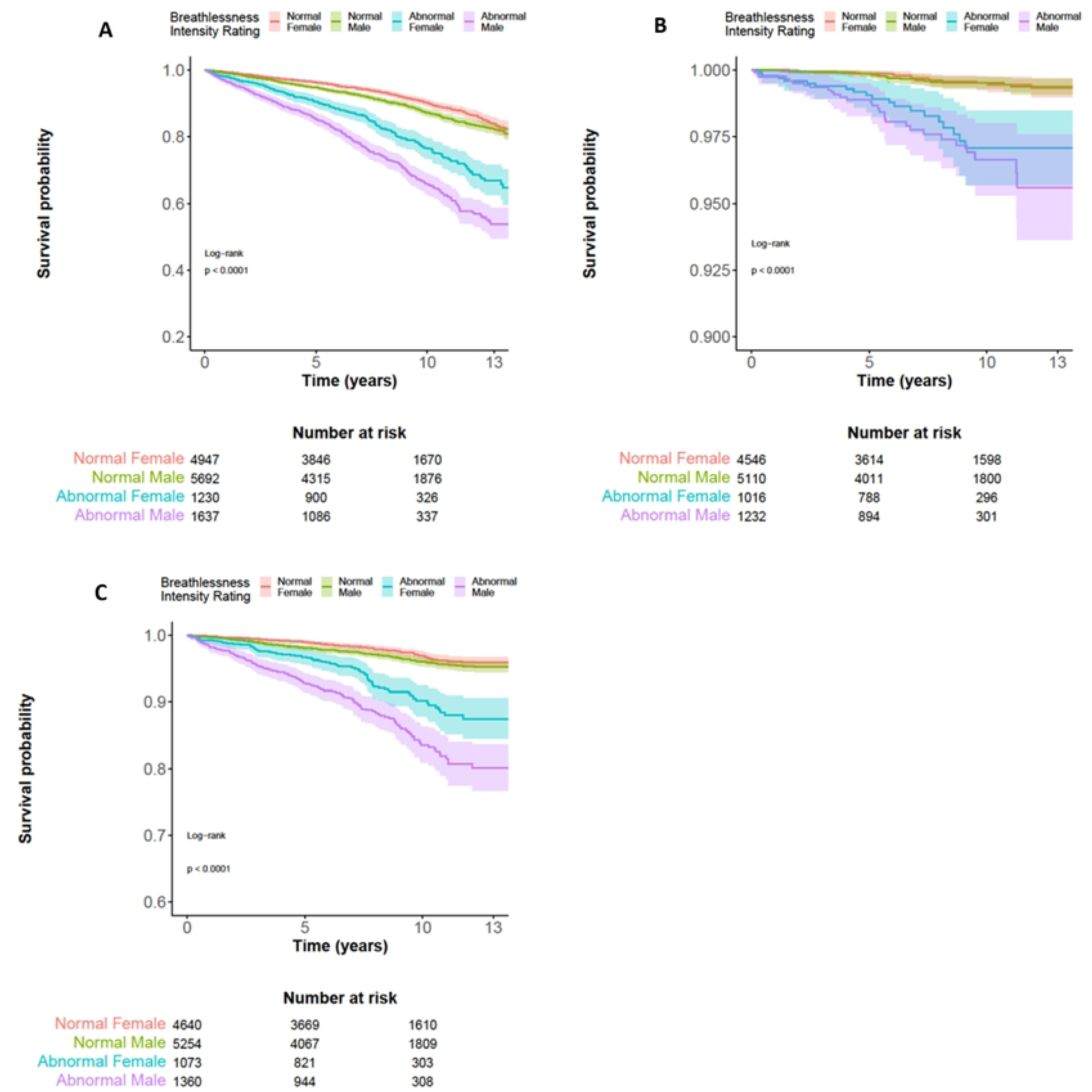
Cumulative survival for normal and abnormally high exertional breathlessness intensity rating(s) during incremental cycle exercise testing (IET) for females and males adjusted for age and body mass index. Mortality (survival) is shown on the y-axis and time (years) on the x-axis. Panels a, b and c show all-cause, respiratory and cardiac mortality, respectively. Abnormally high exertional breathlessness intensity is associated with a higher risk of all-cause, respiratory and cardiac mortality for both males and females (p < 0.001 for both sexes).

**Figure 3.**
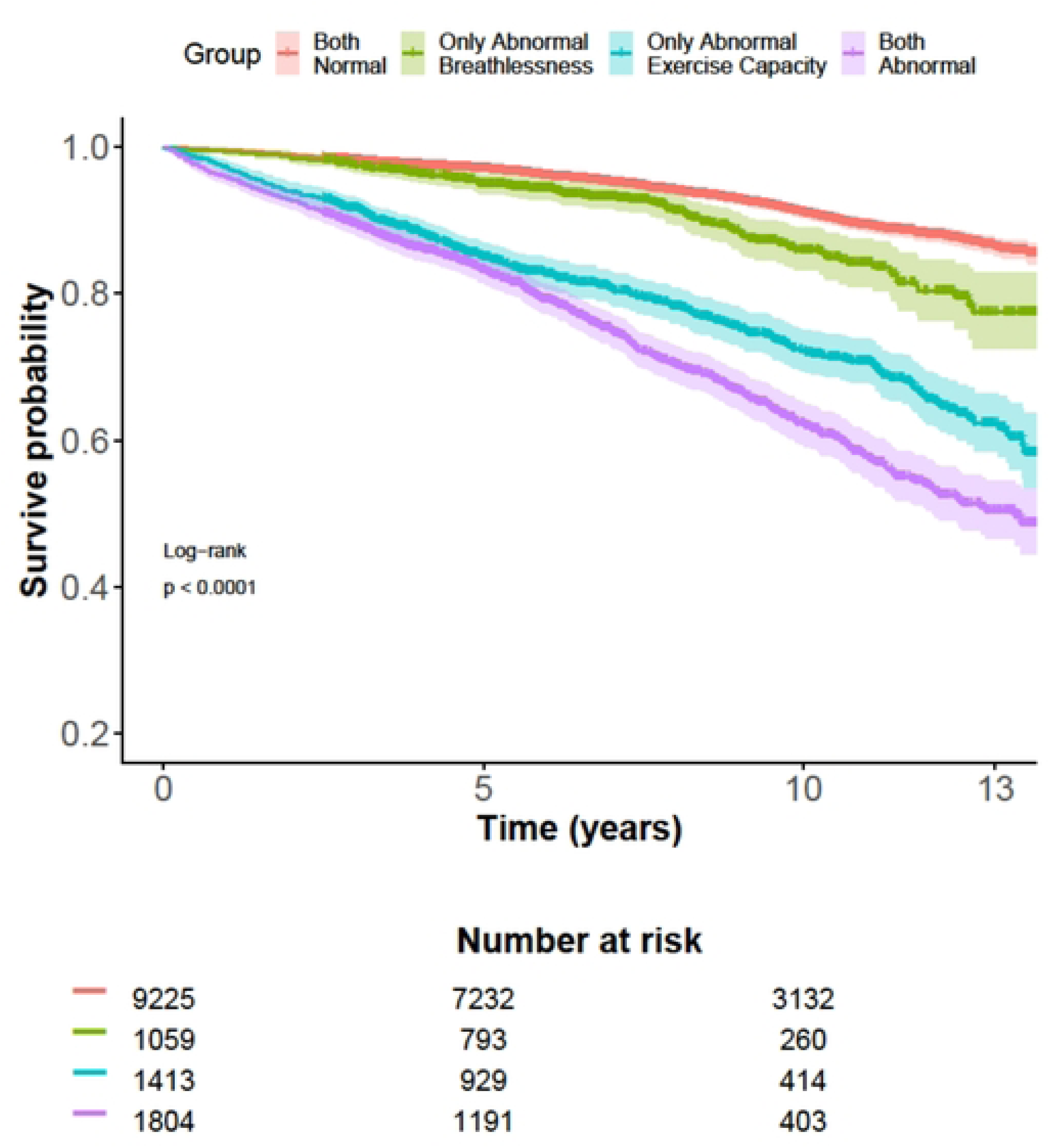
Cumulative all-cause survival grouped by normality of exertional breathlessness and exercise capacity adjusted for age, sex and body mass index. Hazard ratio (HR) and 95% confidence intervals for all-cause mortality separately for abnormally high compared with normal exertional breathlessness intensity. Abnormally low exercise capacity was defined as having a peak exercise capacity below 75% of the predicted normal value. C-statistics showed a discriminatory value of 0.82 (n = 13,494, No. of events = 1,683). Both normal (n = 9,225): HR = 1 (reference) Only abnormal breathlessness (n = 1,059): HR = 1.5 (1.2-1.8) Only abnormal exercise capacity (n = 1,413): HR = 2.7 (2.4-3.1) Both abnormal (n = 1,804): HR = 3.7 (3.3-4.1)

**Table 2.** Associations between abnormally high exertional breathlessness intensity during incremental cycle exercise testing with all-cause, respiratory and cardiac mortality.

|  | N | Events, n (%) | HR (95% CI) | C Statistic |
| --- | --- | --- | --- | --- |
| All-cause | 13,499 | 1,687 | 2.3 (2.1, 2.6) | 0.80 |
| Respiratory mortality | 11,897 | 85 | 5.2 (3.4, 8.0) | 0.87 |
| Cardiac mortality | 12,320 | 508 | 3.0 (2.5, 3.6) | 0.86 |

Hazard ratio (HR) and 95% confidence intervals (CI) for all-cause, respiratory and cardiac mortality separately for abnormally high exertional compared with normal exertional breathlessness intensity. Adjusted by age, body mass index, and sex for the whole cohort. Abnormally high breathlessness was defined as being above the ULN at any one or more exercise stages (all-together).

The association between abnormally high exertional breathlessness and all-cause mortality was similar across all %predW_max_ stages, except at 100% predW_max_ for females, in which there were no deaths in the group with abnormally high breathlessness. The strongest association between abnormally high exertional breathlessness and all-cause mortality was observed at 50% predW_max_ for males (HR = 2.6 [2.2-3.0]) and at 25% predW_max_ for females (HR = 4.8 [2.4-9.4]). The association between all-cause mortality and abnormally high exertional breathlessness intensity for the last measured breathlessness intensity rating during IET was similar to having abnormally high exertional breathlessness at any one or combination %predW_max_ stages (HR = 2.4 [2.1, 2.6]) (Table 3).

**Table 3.** Associations between abnormally high exertional breathlessness intensity ratings at various measurement time points during incremental cycle exercise testing with all-cause mortality.

|  | Exercise intensity (% of predicted Wmax) |  |  |  |  |  |
| --- | --- | --- | --- | --- | --- | --- |
|  | 25% (15 to < 35) | 50% (40 to < 60) | 75% (65 to < 85) | 100% (90 to < 110) | Last measured | All together |
| **Males** | 1,014 | 6,485 | 6,049 | 2,767 | 7,326 | 7,326 |
| Deaths (% deaths) | 76 (7.5%) | 844 (13%) | 642 (11%) | 141 (11%) | 1,036 (14%) | 1,036 (14%) |
| HR (95% CI) | 1.8 (1.1, 3.0) | 2.6 (2.2, 3.0) | 1.8 (1.5, 2.2) | 1.7 (0.8, 3.5) | 2.4 (2.1, 2.7) | 2.4 (2.1, 2.7) |
| C-Statistic | 0.84 | 0.80 | 0.82 | 0.83 | 0.81 | 0.81 |
| **Females** | 690 | 5,511 | 5,168 | 2,635 | 6,173 | 6,173 |
| Deaths (% deaths) | 37 (5%) | 544 (10%) | 442 (9%) | 120 (5%) | 651 (11%) | 651 (11%) |
| HR (95% CI) | 4.8 (2.4, 9.4) | 2.4 (2.0, 3.0) | 1.6 (1.2, 2.0) | NA | 2.2 (1.9, 2.6) | 2.2 (1.8, 2.5) |
| C-Statistic | 0.86 | 0.78 | 0.78 | 0.77 | 0.78 | 0.78 |
| **All** | 1,704 | 11,995 | 11,217 | 5,402 | 13,499 | 13,499 |
| Deaths (% deaths) | 113 (7%) | 1,388 (12%) | 1,084 (10%) | 261 (5%) | 1,687 (12%) | 1,687 (12%) |
| HR (95% CI) | 2.5 (1.7, 3.8) | 2.5 (2.2, 2.8) | 1.7 (1.5, 2.0) | 1.0 (0.5, 2.0) | 2.4 (2.1, 2.6) | 2.3 (2.1, 2.6) |
| C-Statistic | 0.85 | 0.80 | 0.80 | 0.80 | 0.80 | 0.80 |

Hazard ratio (HR) and 95% confidence intervals (CI) of abnormally high compared with normal exertional breathlessness intensity. Analyses adjusted by age, body mass index, and sex for the whole cohort. HR are presented for 25% intervals of W % of predicted max as well as for the last measured breathlessness intensity rating in relation to the %predWmax at that level. The HR for females at 100% is missing. The reason is that there were no death events for the abnormal group.

Participants with normal exercise capacity + abnormally high breathlessness intensity had a HR of 1.5 (1.2-1.8) for all-cause mortality (Group 2). Participants with abnormally low exercise capacity had a HR of 2.7 (2.4-3.1) if normal breathlessness intensity (Group 3), and if abnormal breathlessness intensity a HR of 3.7 (3.3-4.1) (Group 4). C-statistics showed a strong and similar discriminative ability with values of about 0.8 for the evaluated models and %predW_max_ stages (Table 3).

## DISCUSSION

This is the first study to look at the prognostic impact of having an abnormally high breathlessness response to IET, evaluated using standardized reference equations (12) for exertional breathlessness intensity during IET among people in Sweden. The primary finding is that among a large group referred for clinical IET, the presence of abnormally high breathlessness intensity was independently associated with all-cause, cardiac and respiratory mortality. The relative risk of all-cause mortality was approximately 250% in people with compared to without abnormally high exertional breathlessness intensity.

Predictive ability and association between abnormally high exertional breathlessness intensity and all-cause mortality was similar across different %predW_max_ stages, although the number of participants contributing to these analyses naturally decreased as the %predW_max_ stage increased.

Even though the association between breathlessness intensity and mortality was attenuated in the presence of an abnormally low exercise capacity, there remained an independent association. This attenuation is to be expected as exercise capacity is a major determinant of mortality among people performing IET (24).

Based on the present findings, we recommend using a breathlessness intensity rating obtained at peak exercise corresponding to “last measured” in this study. The “last measured” breathlessness intensity rating during IET performed equally well to other exercise stages and “all together” regarding predictive ability and might make application of the reference equations more straightforward as it would be possible to only inquire about breathlessness intensity at the end of test. It would also facilitate comparison of the breathlessness intensity response with other measures (i.e. exercise capacity and other physiological responses) at peak exercise.

As the reference equations always take %predW_max_ into account, a peak breathlessness intensity rating of 6 Borg CR10 scale units, might be abnormal at an abnormally low peak exercise capacity of 40% predW_max_, but within normal limits when a person’s peak exercise capacity is 110% predW_max_. Using the peak recorded value also has the advantage of greater inclusion, as everyone might not reach a specific higher %predW_max_ because of deconditioning or other reasons. Using the peak value thus has the advantage of simplicity, including all people and making comparisons with other measures at peak exercise more straightforward.

The association between abnormally high exertional breathlessness and all-cause mortality is likely driven by several factors. Firstly, in the group with abnormally high compared to normal exertional breathlessness intensity, there was a larger proportion of participants with a health condition(s), such as COPD or HF. Secondly, breathlessness often leads to reduced physical activity ,which in turn can cause a vicious cycle of increased breathlessness and even less physical activity resulting in worsening prognosis (1). There was a trend towards a greater association between respiratory mortality and abnormally high exertional breathlessness as compared to cardiac mortality, although the difference was not statistically significant and might have been influenced by the relatively low number of respiratory-related deaths in this cohort.

Measurement of breathlessness burden is often done using standardized task-based questionnaires (e.g., mMRC dyspnoea scale) in a clinical setting. Gustafsson et al. (28) showed that the mMRC was insensitive to detecting abnormally high exertional breathlessness intensity during cycle IET when evaluated against our recently published reference values for assessing the normality of breathlessness intensity during IET (12). Gustafsson et al. found that a mMRC dyspnoea rating of ≥ 2 only identified 28% of the persons with abnormally high breathlessness during IET, implying that the great majority of people with abnormally high exertional breathlessness may remain undetected if only the mMRC dyspnoea scale was used to quantify breathlessness burden. This strengthens the argument for use of a more standardized test such as IET for detection and evaluation of abnormally high exertional breathlessness as a large proportion of people at risk of premature death might otherwise remain undetected.

Breathlessness intensity ratings during IET can thus be used both to stratify the severity of exertional breathlessness as well as to determine the risk of premature death. From a clinical exercise testing perspective, the results of this analysis suggest that people undergoing a cycle IET do not necessarily need to exert maximum effort to identify abnormally high exertional breathlessness and greater risk of premature death. This could be especially beneficial in people with severe disease or other types of disabilities that preclude maximal exertion.

### Strengths and limitations

Strengths of this study include the large number of people with complete and long follow up (median 8 years) to the hard endpoints of all-cause, respiratory and cardiac mortality using mandatory national registry data.

Physiological data were limited, with no access to pulmonary function tests, ventilation, or gas exchange, limiting our ability to ensure that participants provided maximal effort during the IET (i.e. respiratory exchange ratio). However, mean peak heart rate was about 90% of the predicted maximal value and mean peak Borg RPE ratings were 17-18 implying maximal effort. The present findings pertain to IET performed on a cycle ergometer (non-weight-bearing exercise and are likely not directly applicable to tests performed on a treadmill (weight-bearing exercise) given differences in cardiac, metabolic and ventilatory responses and greater difficulty quantifying power output during treadmill exercise (29, 30).

While the prevalence of some comorbidities, i.e. COPD, were likely underestimated, they were obtained from the National Patient Register which contains all physician diagnosis nationwide coded according to ICD10. This ensured that all who had received a diagnosis were identified correctly.

Further limitations include the lack of a validation cohort as well as the study being conducted at a single center in Sweden with presumably an overwhelming majority of participants being of north European ethnicity. We also had no information on the reasons behind the individual persons abnormally high exertional breathlessness during exercise. We did not have specific information on smoking in the studied group. We know however that the average prevalence of smokers in the Swedish region where the study was done varied between 14% (95% CI: 11–16) and 11% (95% CI: 9–13) during the study period. (31).

### Implications

Abnormally high exertional breathlessness during cycle IET is helpful to identify people at abnormally high risk of all-cause, respiratory and cardiac mortality which could lead to further clinical evaluation. The results could also provide a basis for different types of interventions; for example, cardiopulmonary rehabilitation programs and medical interventions both from a patient and economic perspective. For example, if a person were determined to have abnormally high exertional breathlessness by cycle IET, and thus elevated risk of premature death, further clinical evaluation could be performed with for example cardiopulmonary exercise testing, and other methods to identify the cause. The cause could then be addressed with appropriate intervention.

### Conclusion

Among people referred for clinical exercise testing, the presence of abnormally high breathlessness intensity during cycle IET is associated with all-cause, respiratory and cardiac mortality. Thus, applying reference equations for exertional breathlessness, it is possible not only to determine if abnormal exertional breathlessness is present during IET, but also to establish the associated increase in mortality risk, which we believe has important clinical and research implications.

## Data Availability

As stated by Swedish Ethical Review Authority analysis (Dnr 2018/141-31 ) approving the analysis of this study the data is not allowed to be shared publicly. Sharing sensitive data such as health data publicly is not complying to article 9 of the General Data Protection Regulation (EU 2016/679) as this would compromise the privacy of the participants. The General Data Protection Regulation (EU 2016/679) also considers de-identified sensitive data as sufficient to risk the privacy of participants. According to Swedish law (2003:460) concerning research including humans, ethical permission is required to process data including humans. To access the data from the study, ethical approval first needs to be required from the Swedish Ethical Review Authority(https://etikprovningsmyndigheten.se). Researchers can then contact the corresponding author Viktor Elmberg with suggestions for analysis.

## Acknowledgements and authorship

VE, DJ and ME conceptualized the study; LB collected the data; XZ performed the analyses; VE, XZ, TL, KH, AM, HL, DJ, LB, and ME contributed substantially to the study design, data analysis and interpretation, and the writing of the manuscript.

